# Modeling brain sex in the limbic system as phenotype for female-prevalent mental disorders

**DOI:** 10.1101/2023.08.17.23294165

**Authors:** Gloria Matte Bon, Dominik Kraft, Erika Comasco, Birgit Derntl, Tobias Kaufmann

## Abstract

**Background:** Profound sex differences exist in the prevalence and clinical manifestation of several mental disorders, suggesting that sex-specific brain phenotypes may play key roles. Previous research used machine learning models to classify sex from imaging data of the whole brain and studied the association of class probabilities with mental health, potentially overlooking regional specific characteristics.

**Methods:** We here investigated if a regionally constrained model of brain volumetric imaging data may provide estimates that are more sensitive to mental health than whole brain-based estimates. Given its known role in emotional processing and mood disorders, we focused on the limbic system. Using two different cohorts of healthy subjects, the Human Connectome Project and the Queensland Twin IMaging, we investigated sex differences and heritability of brain volumes of limbic structures compared to non-limbic structures. We applied regionally constrained machine learning models for brain sex classification based solely on limbic or non-limbic features and compared the results with a whole brain model. To investigate the biological underpinnings of such models, we assessed the heritability of the obtained estimates, and we investigated the association with major depression diagnosis in an independent clinical sample.

**Results:** Limbic structures show greater sex differences and are more heritable compared to non-limbic structures. Consequently, machine learning models performed well at classifying sex based solely on limbic structures and achieved performance as high as those on non-limbic or whole brain data, despite the much smaller amount of features in the limbic system. The resulting class probabilities were heritable, suggesting potentially meaningful underlying biological information. Applied to an independent population with major depressive disorder, we found that depression is significantly associated with male-female class probabilities, with largest effects obtained using the limbic model.

**Conclusions:** Overall, our results highlight the potential utility of regionally constrained models of brain sex to better understand the link between sex differences in the brain and mental disorders.

**Highlights:**

- We assessed sex differences and heritability of limbic and non-limbic volumes.
- Limbic volumes showed stronger sex differences and higher heritability overall.
- We trained brain sex classification models on limbic or non-limbic volumes.
- Performance was high and the sex class probabilities were heritable for all models.
- In females, limbic estimates were associated with depression diagnosis.

**Plain English Summary:** Psychiatric disorders have different prevalence between sexes, with women being twice as likely to develop depression and anxiety across the lifespan. Previous studies have investigated sex differences in brain structure that might contribute to this prevalence but have mostly focused on a single-structure level, potentially overlooking the interplay between brain regions. Sex differences in structures responsible for emotional regulation (limbic system), affected in many psychiatric disorders, have been previously reported. Here, we apply a machine learning model to obtain an estimate of brain sex for each participant based on the volumes of multiple brain regions. Particularly, we compared the estimates obtained with a model based solely on limbic structures with those obtained with a non-limbic model (entire brain except limbic structures) and a whole brain model. To investigate the genetic determinants of the models, we assessed the heritability of the estimates between identical twins and fraternal twins. The estimates of all our models were heritable, suggesting a genetic component contributing to brain sex. Finally, to investigate the association with mental health, we compared brain sex estimates in healthy subjects and in a depressed population. We found an association between depression and brain sex in females for the limbic model, but not for the non-limbic model. No effect was found in males. Overall, our results highlight the potential utility of machine learning models of brain sex based on relevant structures to better understand the link between sex differences in the brain and mental disorders.

## Background

Common mental disorders occur at different prevalence rates between sexes (World Health Organization, 2022). In particular, women are twice as likely to develop anxiety and depression across the lifespan compared to men (Kaczkurkin et al., 2019; Pinares-Garcia et al., 2018; Rubinow & Schmidt, 2019; World Health Organization, 2022). This difference arises after puberty (Rubinow & Schmidt, 2019; Slavich & Sacher, 2019), suggesting the involvement of sex-specific factors in the development of such disorders (DeCasien et al., 2022). To identify these factors, neuroimaging studies have investigated sex differences in brain structure and function (Hillerer et al., 2019; S. Liu et al., 2020; Pletzer, 2019; Ritchie et al., 2018; Ruigrok et al., 2014), mostly using univariate analyses. However, discordant findings have been reported (Hillerer et al., 2019; Ritchie et al., 2018; Ruigrok et al., 2014). In a recent meta-analysis, Ritchie et al. (2018) found generally larger volumes in males, while other studies reported larger volumes in females for different regions (S. Liu et al., 2020; Ruigrok et al., 2014). Possible explanations might be differences in the normalization and segmentation processes (Eliot et al., 2021; Hillerer et al., 2019), as well as the effects of total brain volume (Eliot et al., 2021). Sex differences in total brain volume has been shown to drive many structural and volumetric differences in the brain (Dhamala et al., 2022; Eliot et al., 2021), leading to discordant results depending on the correction method applied. In addition, variations in structural and functional MRI according to the menstrual cycle and hormonal contraceptive use have been reported in the literature for many structures, such as hippocampus, amygdala, prefrontal cortex, cingulate cortex, and insula (Dubol et al., 2021; Rehbein et al., 2021). Of note, many of the structures with notable sex differences and hormonal effects are part of the limbic system (Catenaccio et al., 2016; Greve et al., 2021; Grodd et al., 2020; Roxo et al., 2011; Yamagata et al., 2016). The limbic system is strongly involved in emotional processing, learning and memory, functions typically altered in mental and neurological disorders (Lindquist et al., 2012). Due to its involvement in such functions, the limbic system has consequently been proposed as a key player in mood disorders such as major depression (Koolschijn et al., 2009; Sacher et al., 2012; Videbech, 2004; Zheng et al., 2021).

Recently, multivariate approaches have been developed to study sex differences in the brain. Machine learning models that classify for sex based on brain structural or functional magnetic resonance images (MRI) yield class probabilities that can be used as an imaging-derived multivariate phenotype to study sex differences on a continuum from female-to male-like brains (Kim et al., 2022; Tunç et al., 2016; Vosberg et al., 2020; Weis et al., 2020). Conceptually similar approaches have already been used extensively to predict brain age (Cole & Franke, 2017; de Lange et al., 2019, 2020; Franke et al., 2010; Kaufmann et al., 2019; Popescu et al., 2021; Sanford et al., 2022), where machine learning models deliver a continuous phenotype reflecting apparent aging effects. While most brain age studies to date built models based on data from the whole brain, regionally constrained models may identify region-specific associations with mental health, such as frontal brain age alterations in schizophrenia or subcortical alterations in Alzheimer’s disease (Kaufmann et al., 2019). In a recent study, Sanford and colleagues (2022) investigated sex differences in local brain age gaps (i.e. difference between regionally constrained neuroimaging-predicted age and chronological age) in young adults. Compared to males, females showed significantly lower local brain age gap in the frontal region and insula, while they had significantly higher local brain age gap in the posterior regions. However, on a global scale the authors did not report any differences in brain age gaps, suggesting finer-grained (i.e., regional specific) models having a higher sensitivity to sex differences. Translating this finding into the field of sex classification, leveraging regional constrained models may provide estimates more sensitive to sex-specific phenotypes. Whereas Weis and colleagues (2020) have classified sex based on the whole brain connectome and different functional brain networks separately, the potential of regionally constrained models to study brain sex based on structural MRI has yet to be investigated.

Here, we investigate whether a regionally constrained model based on brain volumes of the limbic system can correctly classify sex, and whether the obtained regional class probabilities are sensitive to mental health. We compared limbic brain sex to non-limbic brain sex, aiming to investigate relevant biological differences in brain sex determination as well as the possible clinical association with major depression. Specifically, the present study sought: i) to compare sex differences at a univariate (i.e., single structure) level between limbic and non-limbic structures, ii) to validate regionally constrained machine learning models trained either on limbic or non-limbic feature sets as compared to a whole brain model, iii) to test for an association between obtained class probabilities and major depressive disorder (MDD) diagnosis. We hypothesize that (i) the regionally constrained models, much like whole brain models, are able to correctly predict sex from structural imaging data, (ii) the estimates (i.e., class probabilities) contain biologically meaningful variation (tested via heritability analysis), and that (iii) the limbic estimates have a stronger association with depression than estimates from other models.

## Methods

### Participant selection

As illustrated in Figure 1, structural MRI data from the Human Connectome Project (HCP) (Van Essen et al., 2013) was used for univariate analysis of brain features, and for training of machine learning models in a healthy sample. For the HCP, the subject selection criteria (see Supplementary Figure S1, Additional File 1) aimed to (I) maintain an equal female-male ratio (based on biological sex as provided by the data), while (II) limiting possible confounding effects such as hormonal fluctuation, and (III) maximizing the sample size for machine learning. To achieve this, after excluding 14 individuals following quality control of the imaging data, female subjects were first selected according to the hormonal information available with the goal to limit the potential effects of irregular cycle and hormonal alterations such as presence of hypo- or hyperthyroidism and Thyroid Stimulating Hormone (TSH) levels out of the normal range (0.4-4.0 mU/L, as define by the HCP-YA data dictionary), yielding n = 391 females. These females were then matched to an equal number of males according to age and Euler number. Finally, in order to maximize the sample size, the remaining males (n=105) were matched for age and Euler number with an equal number of randomly selected females from the subjects excluded in the first step. This procedure led to a total sample size of N = 992 subjects (50% females), with an age range 22-38 years old (females: mean age = 28.98, sd = 3.63; males: mean age = 27.90, sd = 3.60).

**Figure 1.**
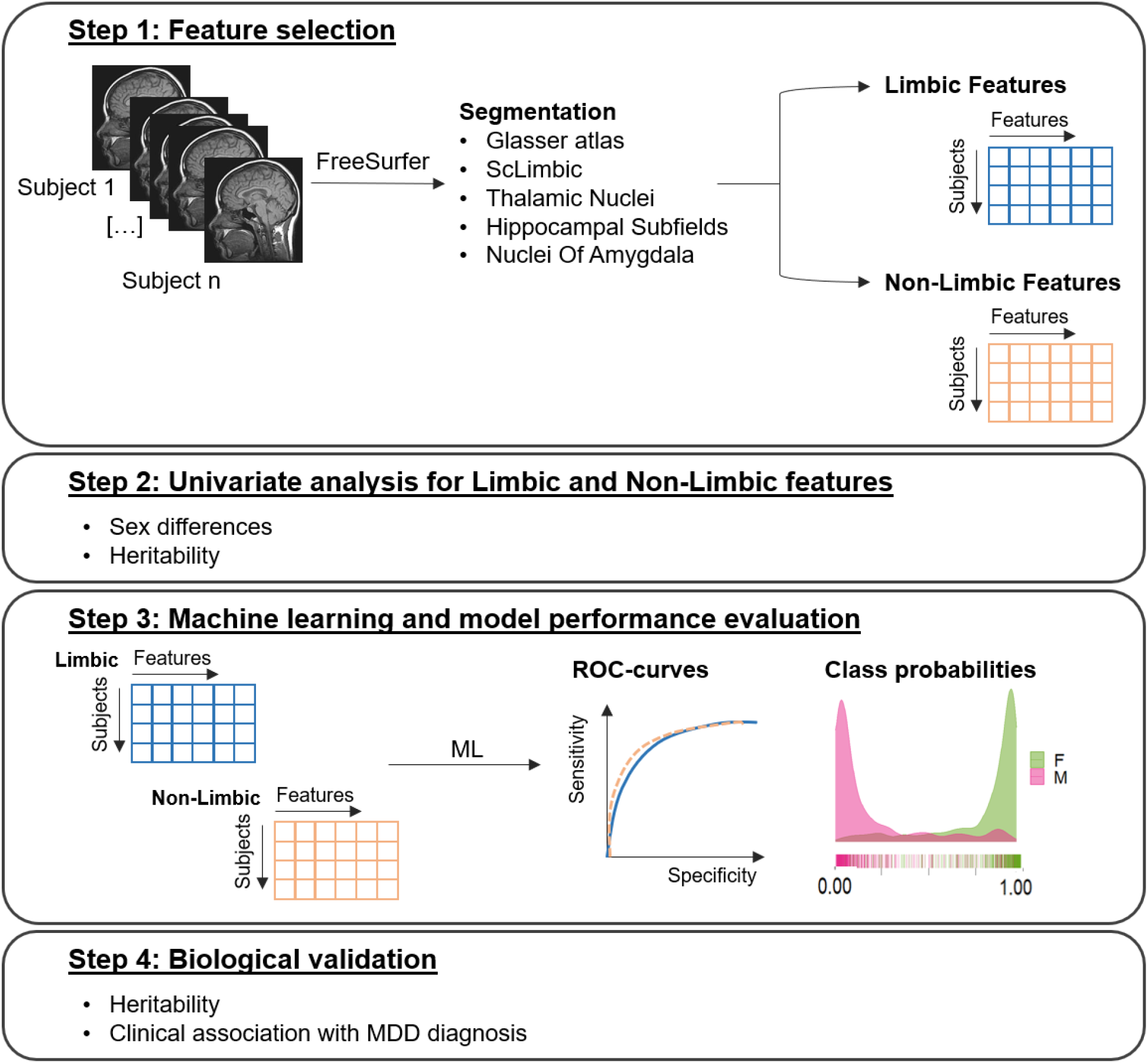
Schematic overview of the study design. <u>Step 1:</u> After combining different segmentations in FreeSurfer, each structure was assigned either to the limbic or to the non-limbic feature set. <u>Step 2:</u> Univariate analysis of sex differences and feature heritability were applied to the limbic and non-limbic feature set. <u>Step 3:</u> A machine learning model for sex classification was trained on each feature set (regionally constrained models). Model performance was assessed via AUC-ROC and the class probabilities for each participant were stored. <u>Step 4:</u> The heritability and the clinical association with major depressive disorder (MDD) diagnosis was assessed for the class probabilities obtained for each model.

The Queensland Twin Imaging (QTIM) study was used as an independent healthy validation dataset (Strike et al., 2022) to replicate univariate analyses and to test the HCP-trained machine learning models. After excluding 20 outliers based on imaging quality control, only subjects in the age range 18-30 years were selected, to ensure a similar age range with the HCP. The final QTIM sample comprised N = 1017 subjects (61.6% females, age range 18-30 years old, females: mean age = 22.49, sd = 2.84; males: mean age = 22.29, sd = 2.86).

To test for clinical associations with the derived brain sex class probabilities, data from the Strategic Research Program for the Promotion of Brain Science (SRPBS) (Tanaka et al., 2021) was used. Due to data availability and the differences in prevalence across sexes (World Health Organization, 2022), we focus on MDD. After excluding 10 outliers following quality control, we selected healthy controls (HC) and individuals with MDD, based on the diagnosis variable available in the dataset. To reduce scan site confounds, we only included HC data for sites in which MDD data was available. This yielded a sample of N = 844 subjects (HC: N = 595, 54.5% female, age range 18-80 years old, females: mean age = 41.34, sd = 14.99, males: mean age = 38.24, sd = 16.28; MDD: N = 249, 47.8% female, age range 18-75 years old, females: mean age = 43.39, sd = 12.87, males: mean age = 41.77, sd = 11.04).

### Image segmentation, quality control and features selection

Raw T1-weighted MRI scans were preprocessed in FreeSurfer v7 and automated cortical and subcortical reconstruction were performed. To obtain a more precise segmentation of the limbic system, we combined volumes obtained with different segmentation approaches. The cerebral cortex was segmented using a multimodal parcellation described in Glasser et al. (2016), which returns 180 features for each hemisphere. For subcortical regions, we combined the classic set of features from FreeSurfer with additional segmentations of subcortical limbic structures (Greve et al., 2021), hippocampus (Iglesias et al., 2015), amygdala (Saygin et al., 2017), and thalamus subfields (Iglesias et al., 2018). For all segmented cortical and subcortical regions, we calculated the corresponding volumes (mm^3^). We then assigned the structures derived from each segmentation process to a limbic or non-limbic feature set based on the common definition of limbic system as found in the literature (Greve et al., 2021; Grodd et al., 2020; Roxo et al., 2011; Yamagata et al., 2016), carefully avoiding overlap between different segmentation approaches. The final feature set comprises 493 regions (358 features from Glasser multimodal segmentation (Glasser et al., 2016), 17 subcortical features from FreeSurfer classical segmentation, 12 features from the subcortical limbic segmentation (Greve et al., 2021), 38 features from hippocampus segmentation (Iglesias et al., 2015), 18 features from the amygdala segmentation (Saygin et al., 2017) and 50 features from thalami subfields segmentation (Iglesias et al., 2018)), of which we assigned 160 regions to the limbic (see Supplementary Table 1, Additional File 1) and 333 regions to the non-limbic feature set.

To correct for differences in head size, all analysis were replicated accounting for the estimated total intracranial volume (eTIV). For each feature we regressed out the eTIV and took the residuals as new eTIV-corrected feature to fit in the machine learning models. As further control, we compared this eTIV residualisation approach to another approach, the power-corrected proportion method (PCP) (D. Liu et al., 2014; Sanchis-Segura et al., 2019), which assumes an exponential relationship between the raw volume and the eTIV. We correlated the features obtained with the two methods (see Supplementary figure S2, Additional File 1), and since they converged on very similar results, we used the eTIV residualisation approach in all eTIV-accounting analyses.

We used Euler numbers, a proxy of image data quality (Rosen et al., 2018) for quality control of the imaging data. We averaged Euler numbers from the left and right hemisphere and excluded subjects with an average Euler number lower than three standards deviations from the sample mean.

### Sex differences and heritability analyses of single brain volumes

We first investigated differences between limbic and non-limbic structures using univariate analyses for each structure of interest. For each region, we tested for sex differences using linear models that accounted for age and Euler number. Subsequently, we assessed the differences in effect size distributions between limbic and non-limbic structures using a t-test. To account for differences in head size, we repeated the same analysis introducing the eTIV as covariate. Next, we computed the broad sense heritability (see *Heritability analysis*) of volumes for each region and subsequently tested for heritability differences between limbic and non-limbic structures using a t-test.

### Sex classification models and clinical associations

For each set of features (limbic, non-limbic and whole brain), two sex classification models were fitted, one using the raw volumes, and one using the same features after regressing out the intracranial volume. To avoid confounding factors, our training sample was balanced for sex and subjects were matched for age and image quality. We trained our models on HCP data, using gradient tree boosting as implemented in the *xgboost* package in R (version 4.2.2). The learning rate was set at η=0.01 and the initial number of rounds to 1000, and we performed 5-fold cross validation within the training sample. For each iteration the prediction error was assessed and used to determine the optimal iteration number, used to train the final models on the full set of data. We then applied these models to the test samples to classify brain sex. The resulting class probabilities were extracted and used for further analysis. To evaluate each model performance, both the accuracy and the area under the receiving operating characteristic curves (AUC) were calculated.

To test the statistical significance of the models, we performed permutation testing. For each raw and eTIV-controlled set of features, the classification labels (sex of the participants) were randomly permuted 5000 times, while maintaining the feature sets unchanged, resulting in the randomized association between the feature matrix and the labels. For each permutation, 5-fold cross validation was applied. The accuracies were stored and used to build a null distribution that was then compared against the accuracies of the true models. Both a cut-off of 50% (chance level) for the accuracy and AUC and significant statistical results in the permutation testing were considered to evaluate the model as successful in classifying sex.

For external validation, we applied the HCP-trained model to independent data from the QTIM and SBRPS samples and computed the class probabilities in these unseen datasets. In SBRPS data, we tested for clinical association with major depression using a linear model considering the diagnosis as independent variable, class probability obtained with each model as the dependent variable and accounting for sex, age, Euler number and site, as follow:

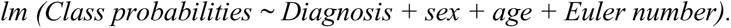

To overcome potential issues with a continuum of class probabilities originating from two distinct distributions (males, females), we repeated the same analysis within each sex. As further control, we repeated the same analyses (general and within each sex) on a subset of subjects with an equal number of HC and MDD subjects matched by age and sex (HC: N = 249; MDD: N = 249; females = 48%). Effect sizes were assessed using Cohen’s d (Cohen, 1988).

### Heritability analysis

Monozygotic and dizygotic twin couples were selected for the HCP and QTIM data. Only couples of twins with both twins in the data were selected in both HCP and QTIM dataset. Due to the sample, in the HCP all included dizygotic couples were of the same sex. In the QTIM sample, both same sex and discordant sex couples of twins were selected. Siblings were excluded from the analysis in both datasets. The total sample sizes for heritability analysis were N = 378 (MZ = 236, DZ = 142) for HCP and N = 674 (MZ = 302, DZ = 372) for QTIM data. The heritability analyses were run at a single-structure level and on the predicted class probabilities from the machine learning models. An AE model was used for both, returning the variance explained by the additive genetic component (A) and non-shared environment component (E). Sex, age and Euler number were accounted as covariates. For the single feature analyses, we repeated the same analysis on the raw volumes and on the residuals after regressing out the covariates and the total intracranial volume to control for head size. The heritability analyses were carried out with the *twinlm* function of the *mets* package in R (version 1.3.1).

## Results

### Limbic structures showed greater sex differences and heritability than non-limbic structures

The degree of sex differences of a given FreeSurfer-derived feature to its heritability in two independent samples (HCP and QTIM) is depicted in Figure 2. Limbic structures showed significantly greater sex differences as compared to non-limbic structures in both the HCP (t = 4.89, p < 0.001) and QTIM (t = 3.87, p < 0.001) data. These results were replicated when accounting for the eTIV in both samples (HCP: t = 5.72, p < 0.001; QTIM: t = 5.59, p < 0.001), suggesting overall greater sex differences independent of head size in the limbic system. The limbic volumes were also more heritable than the non-limbic volumes, both with and without controlling for eTIV, in HCP (Raw features: t = 4.85, p < 0.001; eTIV-controlled: t = 4.84, p < 0.001) and QTIM sample (raw features: t = 3.36, p < 0.001, eTIV-controlled: t = 3.75, p < 0.001).

**Figure 2.**
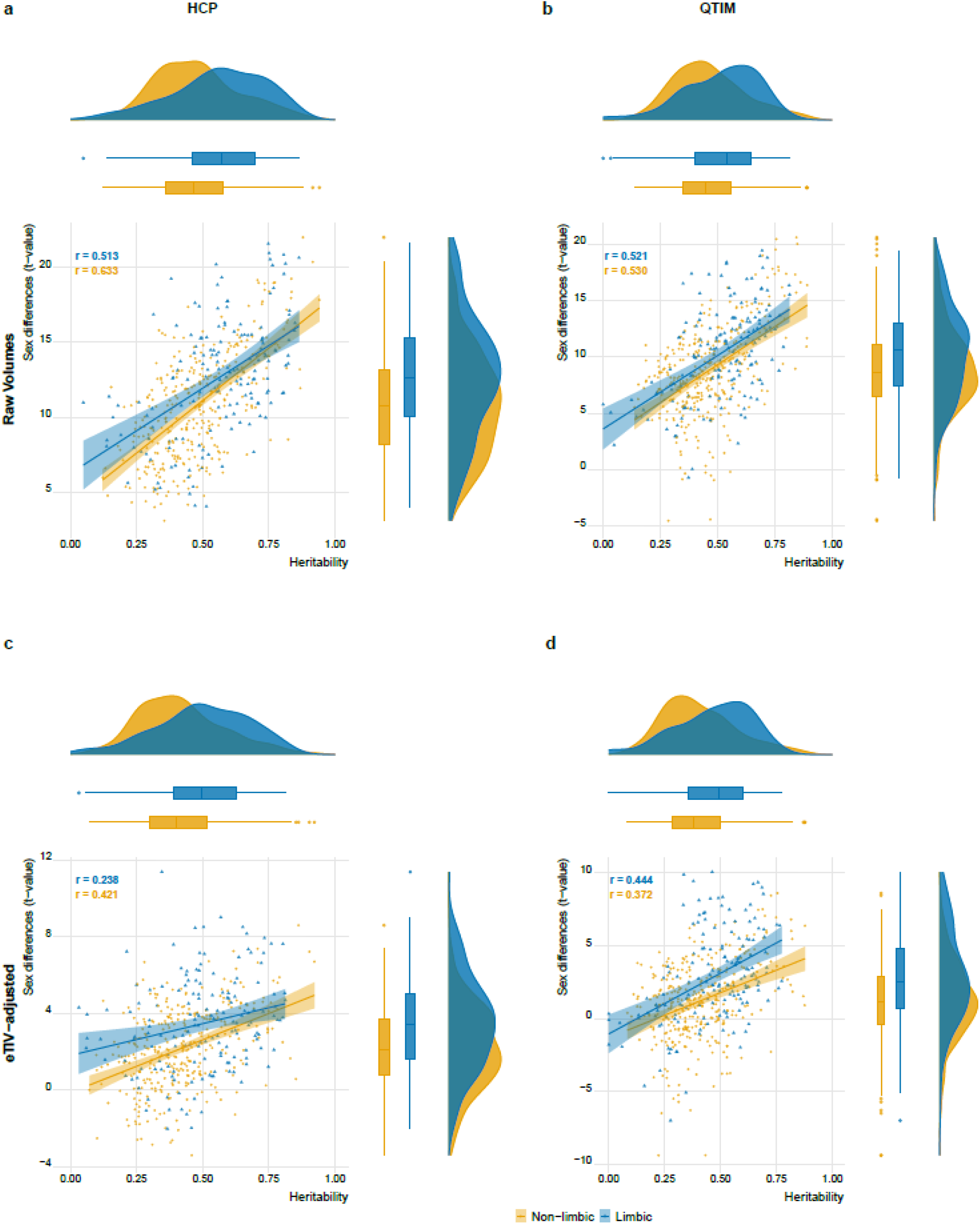
Limbic volumes show stronger sex differences and higher heritability than non-limbic volumes. The analysis was initially conducted in HCP (panel **a, c**) and thereafter replicated in the independent QTIM sample (panel **b, d**). The upper row (**a, b**) presents results from raw volumes whereas the lower row (**c, d**) shows results after controlling for estimated total intracranial volume. Each data point reflects the effect of one brain region. Regression lines illustrate the direction of effect across all limbic or non-limbic regions, indicating that the most heritable features had also grater sex differences. In both samples, the identified effects from raw volumes were smaller yet still significant when accounting for the total intracranial volume.

For both, limbic and non-limbic structures, we observed a significant positive association between sex difference and heritability, indicating that the most heritable features had also greater sex-differences (range across the eight regression lines depicted in Figure 2: r = .238, p = .003 to r = .633 p < 2.2e-16) (see Supplementary Table 2, Additional File 2, for the complete list of values).

### Regionally constrained models successfully classify sex

We used the different feature sets (limbic, non-limbic, whole brain) from the HCP sample to train a machine learning model that classified sex from volumetric imaging data. We first assessed model performance within HCP using 5-fold cross-validation, indicating that all three models were able to successfully classify sex (please see Figure 3 for illustration). The limbic model achieved an accuracy of 87% and AUC of 0.935, while the non-limbic achieved an accuracy of 84.4% and AUC of 0.92. Both models performed approximately as well as the whole brain model (accuracy = 86.6%, AUC = 0.941). When controlling for eTIV, the models were still capable to correctly classify sex, although with a lower accuracy and AUC (limbic (eTIV): accuracy = 70.9%, AUC = 0.778; non-limbic (eTIV): accuracy = 74.6%, AUC = 0.819; whole brain (eTIV): accuracy = 77.2%, AUC = 0.861). No significant difference in the performance between the three models was found.

**Figure 3.**
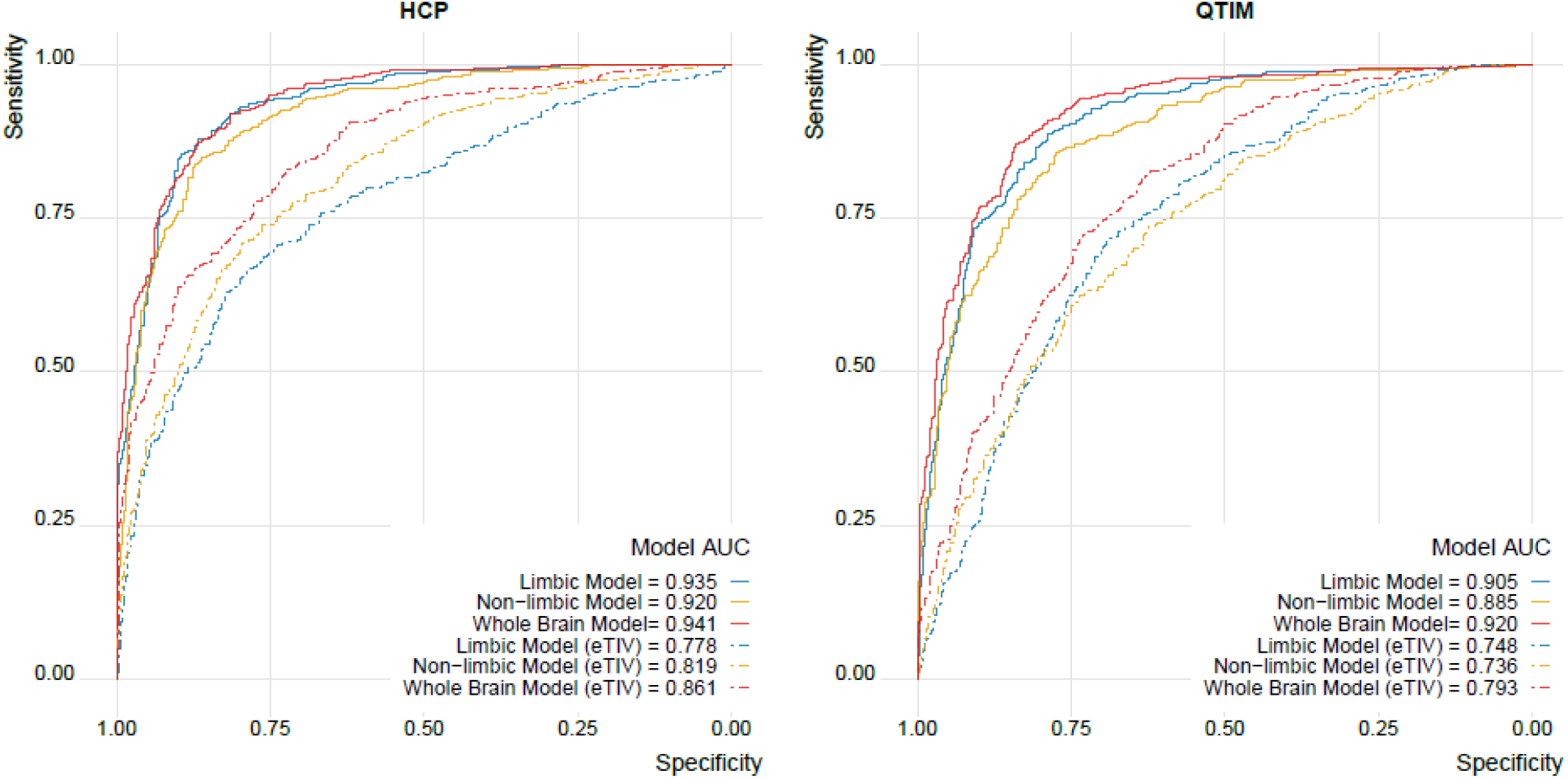
The limbic model achieved performances similar to models based on non-limbic and whole brain data. The ROC curves and the AUC for each model in the HCP sample (cross-validation within the training set) and in the QTIM sample (validation in independent test data) show high performances for all models. Regressing eTIV from the features (dashed lines) reduced performance compared to raw volumes (solid lines) yet all models still performed well above chance.

Permutation testing with 5000 permutations per model indicated that no permutation-based accuracy was better than the accuracy obtained with the true models, confirming the validity of our models (see Supplementary Figure S3, Additional File 1). Notably, when looking into the feature importance for the raw and eTIV-corrected whole brain models, the main contributors to both models belong to the limbic system (see Supplementary Figure S4, Additional File 1).

Next, we applied the HCP-trained models to QTIM data for external validation, confirming solid performance in independent data (with an accuracy of 82.3%, 77.6% and 79.7% and an AUC of 0.905, 0.885 and 0.920 for limbic, non-limbic and whole brain model, respectively). For eTIV accounted models we also observed performances similar to those obtained within HCP (accuracy: 68.4%, 66.8%, 71.6%, AUC: 0.748, 0.736, 0.793 AUC for limbic, non-limbic and whole brain model, respectively).

### Brain sex class probabilities are heritable

The results of the broad sense heritability estimated from twin data of the class probabilities obtained from each of the machine learning models is shown in Figure 4. Sex class probabilities were heritable for all models, both in the HCP (limbic: 81.4%, non-limbic: 89.4%, whole brain: 87.4%) and the QTIM data (limbic: 78.9 %, non-limbic: 82.1%, whole brain: 74.7%). When accounting for the total intracranial volume, the broad sense heritability decreased in both samples yet was still substantial (HCP: limbic (eTIV): 61.3%, non-limbic (eTIV): 47.6%, whole brain (eTIV): 51.98%; QTIM: limbic (eTIV): 48.5%, non-limbic (eTIV): 56.5%, whole brain (eTIV): 57.8%).

**Figure 4.**
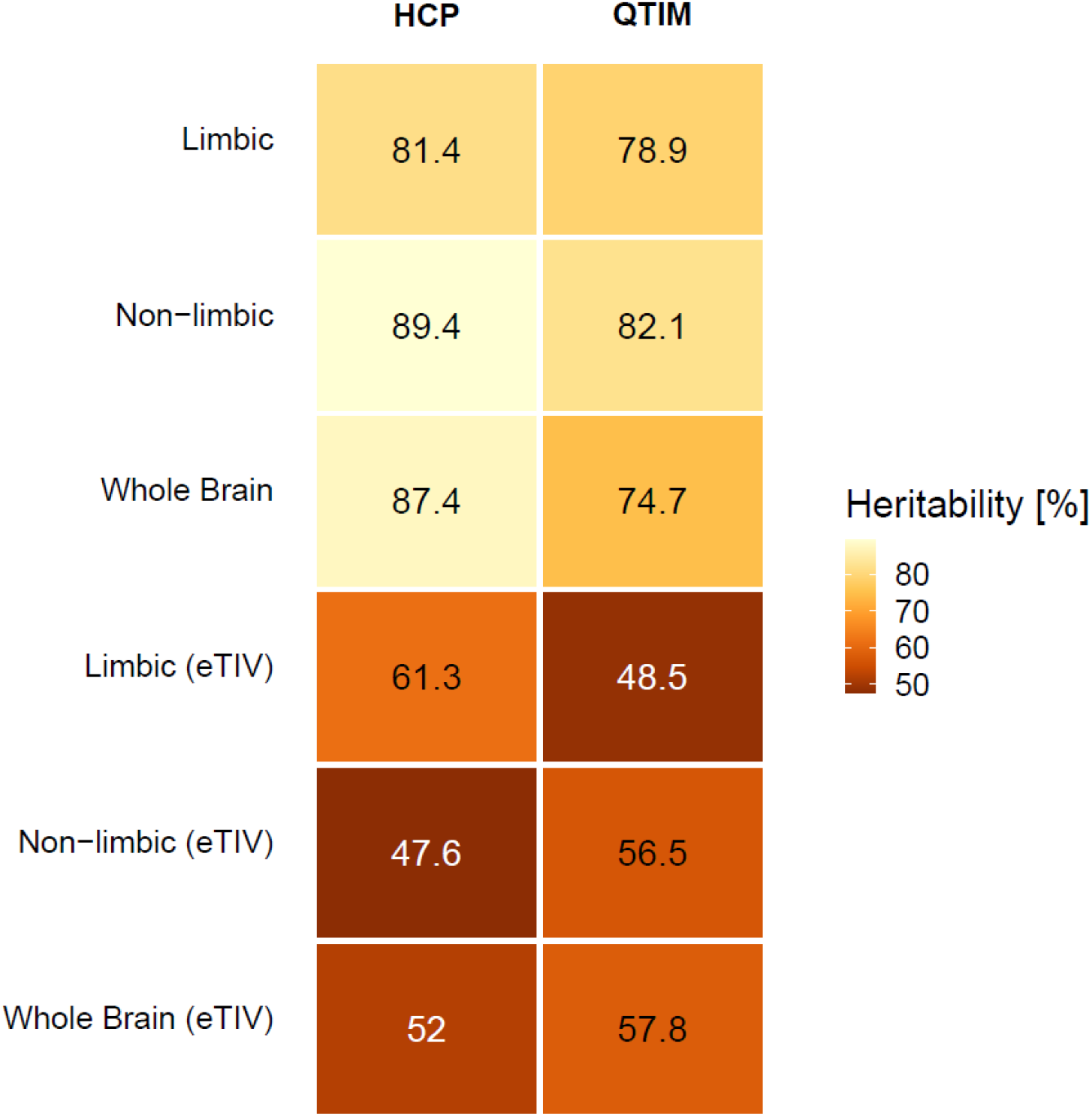
The class probabilities from all models were heritable in both samples, indicating meaningful underlying biological information. Each cell shows the broad sense heritability in percent, with highest values for those derived from models that do not account for eTIV. Similar values are obtained when repeating the analyses within each sex (see Supplementary Figure S5, Additional File 1).

### Higher class probabilities are associated with major depression

To investigate the association with MDD in a clinical sample, we applied the models to the SRPBS sample, considering healthy controls and depressed patients. After verifying the accuracy and the AUC of each model (see Supplementary Figure S6, Additional File 1), we extracted the class probabilities and associated them with the diagnosis. Our results indicated that the class probabilities in the clinical sample were higher (i.e. in the direction of a female brain phenotype) as compared to the healthy controls. These results were significant in all models, with strongest effect for the limbic model (limbic: t-value = 2.81, p = 0.005, Cohen’s d = 0.21; non-limbic: t = 2.57, p = 0.011, Cohen’s d = 0.2; whole: t = 2.40 p = 0.016, Cohen’s d = 0.18). When accounting for eTIV, these results were no longer significant (please see Figure 5 for details). Interestingly, when analyzing the association between class probabilities and depression within sex, no effect was found in males, while only the limbic (t = 3.11, p = 0.002, Cohen’s d = 0.34) and whole brain (t = 2.27, p = 0.023, Cohen’s d = 0.25) models showed significant differences between HC and MDD in females. When accounting for eTIV, none of the associations within sex were significant. However, similar patterns of stronger effects in females compared to males was found for the three models (females: limbic (eTIV): t = 1.94, p = 0.053, Cohen’s d = 0.21; non-limbic (e-TIV): t = 0.15, p = 0.881, Cohen’s d = 0.02; whole brain (eTIV): t = 1.57, p = 0.118, Cohen’s d = 0.17; males: limbic (eTIV): t = 0.13, p = 0.895, Cohen’s d = 0.01; non-limbic (eTIV): t = 1.20, p = 0.231, Cohen’s d = 0.13; whole brain (eTIV): t = 0.27, p = 0.787, Cohen’s d = 0.03). When repeating the analyses in the age-matched HC-MDD subsample, only the limbic model in females showed significant greater class probabilities in MDD (t = 2.14, p = 0.033, Cohen’s d = 0.28), while no significant association was found for the general sample or males (see Supplementary Figure S7, Additional File 1).

**Figure 5.**
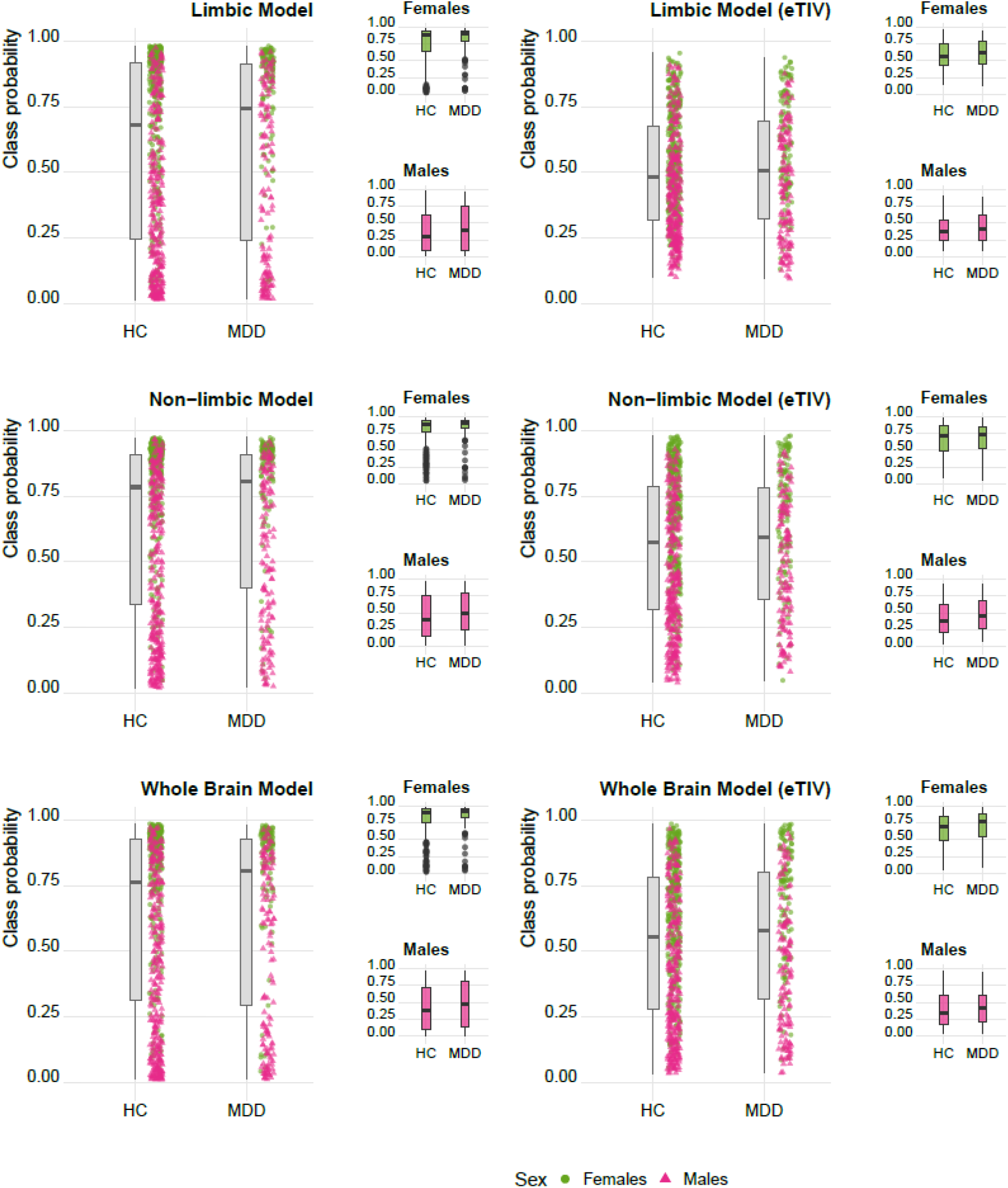
MDD diagnosis is associated with a more female-like brain. Associations between class probabilities and diagnosis for major depression disorders. Higher class probabilities are associated with MDD diagnosis for all raw models. When accounting for eTIV these effects are no longer significant.

## Discussion

The present study aimed at investigating whether regionally constrained machine learning models can correctly classify sex from T1-based volumetric MRI data, investigating if regional class probabilities are more sensitive to a female-prevalent mental disorder compared to whole brain models, the currently used standard in the field. Known for its involvement in various emotional and cognitive abilities as well as its central role for mental conditions, we here focused on the limbic system to investigate brain sex as a putative phenotype to study female-prevalent mental disorders. Our results based on univariate analysis indicate that limbic structures show substantially greater sex differences compared to non-limbic structures. Limbic structures were also more heritable, which may indirectly suggest that their sex differences are mediated by genetic components. These univariate findings supported the utility of the limbic system as a target for regional constrained brain sex prediction. Our multivariate machine learning models using limbic, non-limbic and whole brain features, respectively, were able to classify sex, yielding heritable class probabilities that were associated with major depression diagnosis, suggesting meaningful underlying biological variance.

### Limbic structures showed greater sex differences and heritability than non-limbic structures

In line with previous studies showing associations of limbic structures with sex and hormonal status (Dubol et al., 2021; Hillerer et al., 2019; Pletzer, 2019; Rehbein et al., 2021), our univariate analyses showed that limbic structures were characterized by stronger sex differences compared to non-limbic structures. Likewise, we found stronger heritability for limbic compared to non-limbic structures, which resembles a previous report of high heritability in several limbic structures, including hippocampus, amygdala, and nucleus accumbens (den Braber et al., 2013), and contributes to the global efforts to disentangle genetic contribution to brain structure and function (Adhikari et al., 2018; Batouli et al., 2014; Chiang et al., 2011; den Braber et al., 2013; Kochunov et al., 2015; Lukies et al., 2017). The degree to which genetic contributions to the anatomy of the limbic system are influenced by sex is an understudied topic with mixed results thus far. While some studies find no sex differences in brain volumes (den Braber et al., 2013), others report lower heritability in females, suggesting as possible explanation a greater influence of environmental factors such as the hormonal status in females (Batouli et al., 2014; Lukies et al., 2017). Supporting the latter hypothesis, many limbic structures express receptors for sex hormones (Rehbein et al., 2021; Sundström-Poromaa et al., 2020), contributing to plasticity mechanisms and structural changes. Here, we however found the same pattern of stronger heritability in limbic structure compared to non-limbic structures in males and females, indicating that the heritability patterns were not driven by one sex. Notably though, our results indicate a strong positive association between sex differences and heritability of brain structure, indicating that the structures showing greater sex differences are also the most genetically determined structures, even after controlling for head size.

### Regionally constrained models successfully classify sex

The direct comparison and interpretation of sex effects on specific brain regions using univariate frameworks may be limited by the diversity in terms of protocols and parameters used by different studies (Hillerer et al., 2019; Rehbein et al., 2021). Thus, multivariate analysis in a machine learning framework can provide the advantage of condensing the information from a set of brain features into a single score (the class probability), which could be used as variable for further analysis avoiding difficult comparisons. Here, we demonstrated that sex can be classified from regionally constrained feature sets without performance loss. In particular, our limbic and non-limbic models were both able to classify brain sex with accuracies and AUC similar to those obtained with the whole brain model. It is worth noting that the limbic model achieved descriptively the same level of accuracy and AUC with much less features than the other models (limbic: 160 features, non-limbic: 333, whole brain: 493). It must be noted that the cross-validation procedure implemented in HCP did not account for the presence of family members in the splitting procedure, thus potentially biasing the classification performance via twin similarity. However, the external validation in two independent samples achieved high accuracies for all models, suggesting that the presence of twins in the model training did not induce substantial bias.

### Brain sex class probabilities are heritable

Since deviations in class assignment can reflect both methodological error or biological variance we assessed the degree to which the class probabilities returned by each of the models capture biologically meaningful variance, by first assessing their heritability. Although previous work has investigated heritability of different structural and functional brain measures using univariate analyses (Adhikari et al., 2018; Batouli et al., 2014; Kochunov et al., 2015; Lukies et al., 2017), heritability studies of multivariate estimates of brain sex are scarce. A recent study obtained heritable sex scores from a classification of whole brain data (van Eijk et al., 2021), in line with our results. Here, we extend these findings to regional constrained estimates of brain sex. We found that the class probabilities were heritable for all models, including those controlling for total intracranial volume, supporting their interpretability by pointing at underlying genetic factors.

### Higher class probabilities are associated with major depression

Building upon the heritability results indicating biological meaningful variance, we furthermore investigated the clinical association with major depression under the hypothesis that class probabilities of limbic features may serve as a putative phenotype for the investigation of sex-prevalent mental health conditions. Based on data-availability, we focused on MDD, known to be more prevalent in females (World Health Organization, 2022). Previous univariate analyses have shown alterations in neuroimaging phenotypes in MDD (Grieve et al., 2013; Harris et al., 2022; Koolschijn et al., 2009; Sacher et al., 2012; Schmaal et al., 2016; Videbech, 2004; Zheng et al., 2021), with particular focus on the limbic system. Studies investigating structural changes in brain MRI have displayed reduction in brain volume of several limbic regions in depressed subjects, underling the possible involvement of the limbic system in onset and maintenance of depression (Koolschijn et al., 2009; Sacher et al., 2012; Schmaal et al., 2016; Videbech, 2004; Zheng et al., 2021). However, many of these studies do not account for possible sex differences in the effects, reporting an overall smaller volume of cortical and subcortical structures in MDD patients compared to HC (Koolschijn et al., 2009; Videbech, 2004; Zheng et al., 2021). Multivariate classification models deliver probabilities at the single subject level, facilitating the investigation of sex specific effects in disorder associations. Previous studies attempted to associate brain sex estimates with other common mental disorders and symptoms with sex deviant prevalence (Phillips et al., 2019; van Eijk & Zietsch, 2021). However, these studies only focused on whole brain estimates. We extend this by showing that in all three models (limbic, non-limbic, whole brain) a more female-like brain is associated with MDD diagnosis. Interestingly, the association was strongest for the limbic model, complementing previous univariate findings on the relevance of the limbic system and supporting that regionally constrained estimates might represent sensitive markers to study brain – mental health associations. Moreover, when analyzing the two sexes separately, the effect survived only in females and only in the limbic and whole brain model, highlighting the importance of investigating sex differences in clinical associations with brain sex. Nevertheless, it must be noticed that when controlling for eTIV none of the models were significantly associated with depression diagnosis. Thus, from the data at hand we cannot rule out that the observed clinical associations were driven by sex differences in total brain size.

### Methodological considerations and future directions

Potential limitations may stem from the fact that hormonal status, by acting on brain plasticity, might affect brain structure. Thus, the class probabilities obtained with our model might change according to the phase of the menstrual cycle, the intake of hormonal contraceptives, and the age-related hormonal status of the participants. As noted, many limbic structures are particularly sensitive to these changes (Barth et al., 2016; Dubol et al., 2021; Rehbein et al., 2021). We attempted to mitigate the hormonal effects by matching the subjects in the training data based on the available menstrual cycle information. However, the lack of precise data on the hormonal status of the participants hinders the effort to control for the variability due to hormonal effects. Women’s health factors such as menstrual cycle, hormonal contraceptives use, pregnancy, and menopause are still largely overlooked in neuroimaging research (Taylor et al., 2021). Thus, further neuroimaging data and research considering hormonal levels across the menstrual cycle and different life stages is needed to provide more insights on brain sex classification models and to move the field forward. Moreover, while we placed strong emphasis on replicating our results in independent data, lending credibility in both the univariate analyses (heritability, sex differences) as well as in the multivariate analysis (model performance, heritability of class probability), data availability limited us in the replication of clinical associations. We thus deem it important to validate our clinical associations in another dedicated study, which would also test our results generalizability to different technical and clinical characteristics, such as differences in scan protocols or confounds due to the known impact of antidepressant treatment on brain plasticity (Koolschijn et al., 2009; Schmaal et al., 2016). Finally, although the effects were in the same direction, when controlling for the eTIV our estimates failed to reach the statistical threshold for the clinical association with MDD. Sex differences in total brain volume have been previously associated with structural differences in regional volume (Dhamala et al., 2022; Eliot et al., 2021; Sanchis-Segura et al., 2019, 2020). Recent studies showed how different corrections methods for total intracranial volume might lead to different results in both univariate and multivariate analyses for sex differences (Sanchis-Segura et al., 2019, 2020). Here, we corrected our analysis by regressing out the eTIV from each feature. To further control, we applied the power-corrected proportion approach and correlated it with the results of the regression methods, obtaining a correlation of r > 0.98 for all our features. These results suggest that our eTIV correction is robust (see Supplementary Figure S2, Additional File 1), however, other methods beyond those tested could still yield different results (Sanchis-Segura et al., 2019, 2020).

## Conclusion

In conclusion, we here show in two independent data sets that sex can be classified from T1-based MRI volumes using regionally constrained models, by integrating prior knowledge into the selection of machine learning model features. Our limbic model achieved as high accuracy as the whole brain model using only one third of the features of the latter and the respective class probabilities displayed the strongest associations with major depression diagnosis. Heritability analysis further supports that these probabilities capture biologically meaningful information.

## Perspectives and Significance

Previous studies have deployed machine learning models to classify sex from brain imaging data and derived male-female class probabilities at the individual level. However, the degree to which these probabilities vary across subsets of brain regions remains largely unexplored. Here, we study sex differences in limbic vs. non-limbic brain features and found strongest association of limbic sex probabilities with clinical data. These findings highlight the potential utility of regionally constrained models to investigate the link between brain sex and mental disorders and call for future investigations into other mental disorders with sex differences in prevalence and symptom profiles.

## Data Availability

All data used in the present study are available under data user agreement with the corresponding institutions. HCP data were provided by the Human Connectome Project, WU-Minn Consortium. Queensland Twin IMaging data were available online at https://openneuro.org/datasets/ds004169/versions/1.0.4. The Strategic Research Program for Brain for the Promotion of Brain Science were provided by the DecNef Department at the Advanced Telecommunication Research Institute International, Kyoto, Japan.

## Declarations

### Ethics approval

The HCP study obtained the ethical approvement of the IRB of Washington University. The QTIM study was approved by the Human Research Ethics Committee at the QIMR Berghofer Medical Research Institute (Ref#P701). All participants including a parent or guardian for those aged under 18 years provided written informed consent. The SRPBS was conducted in accordance with the Declaration of Helsinki and the study has been approved by the institutional review boards for each contributing institution. Written informed consent was obtained for all participants (Tanaka et al., 2021).

### Consent for publication

Written informed consent was obtained by each study providing the data.

## Acknowledgements

This work was performed as part of the International Research Training Group: Women’s Mental Health Across the Reproductive Years (IRTG 2804). The study was supported by the BMBF-funded de.NBI Cloud within the German Network for Bioinformatics Infrastructure (de.NBI) (031A537B, 031A533A, 031A538A, 031A533B, 031A535A, 031A537C, 031A534A, 031A532B).

## Availability of data and materials

HCP data were provided [in part] by the Human Connectome Project, WU-Minn Consortium (Principal Investigators: David Van Essen and Kamil Ugurbil; 1U54MH091657) funded by the 16 NIH Institutes and Centers that support the NIH Blueprint for Neuroscience Research; and by the McDonnell Center for Systems Neuroscience at Washington University. In addition, the authors used data from the Queensland Twin IMaging (QTIM, https://openneuro.org/datasets/ds004169/versions/1.0.4) and the Strategic Research Program for Brain for the Promotion of Brain Science (SRPBS, https://bicr-resource.atr.jp/srpbs1600/). Data collection and sharing for the SRPBS data was provided by the DecNef Department at the Advanced Telecommunication Research Institute International, Kyoto, Japan.

Analysis code will be made publicly available after approval of the editorial board.

## Author contribution

**Gloria Matte Bon:** Conceptualization, Formal analysis, Data Curation, Data Interpretation, Visualization, Writing – Original Draft, Writing -Review & Editing. **Dominik Kraft:** Data Interpretation, Writing – Original Draft, Writing – Review & Editing. **Erika Comasco:** Supervision, Funding Acquisition, Writing – Review & Editing. **Birgit Derntl:** Supervision, Funding Acquisition, Writing – Review & Editing. **Tobias Kaufmann:** Conceptualization, Data Curation, Data Interpretation, Supervision, Funding Acquisition, Writing – Original Draft, Writing -Review & Editing.

## Supplementary information

<u>Additional File 1:</u> Supplementary Figures S1-S7, Supplementary Table 1.

<u>Additional File 2:</u> Supplementary Table 2.

