## Additional_file_1 for "Modeling brain sex in the limbic system as phenotype for female-prevalent mental disorders"

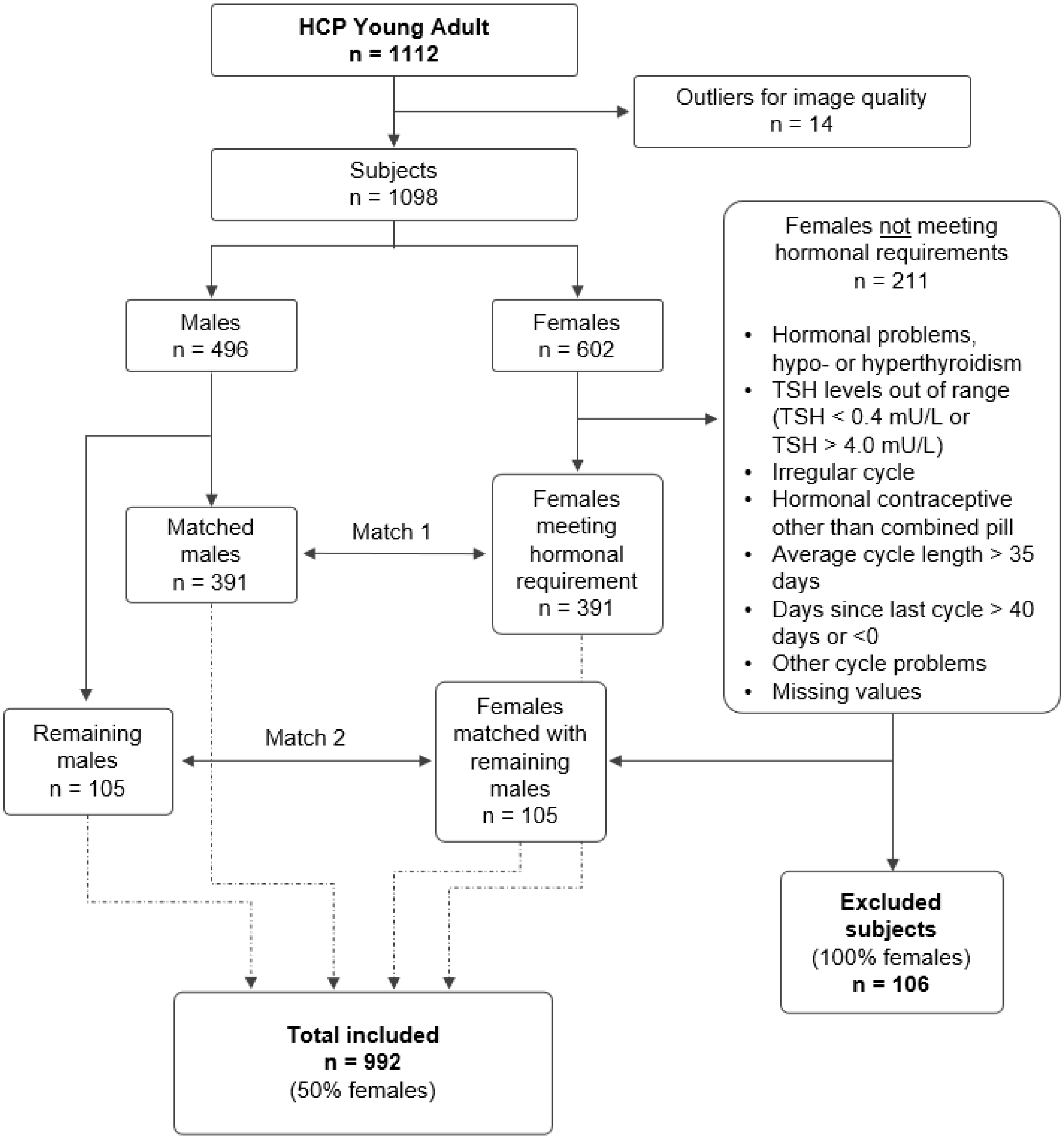


**Supplementary Figure S1. Double matching procedure for Human Connectome Project (HCP) sample.** After excluding outliers for image quality, female subjects were divided in two groups based on the hormonal information available. The group meeting the hormonal requirements was first matched according to age and image quality to a subgroup of males, to limit as much as possible the effects of hormonal fluctuation. Finally, to maximize the sample size, a second matching was applied between the remaining males and a subgroup of females not meeting the hormonal requirements. The final total sample size was N = 992 subjects, with an equal ratio between sexes. Abbreviations: TSH: Thyroid Stimulating Hormone

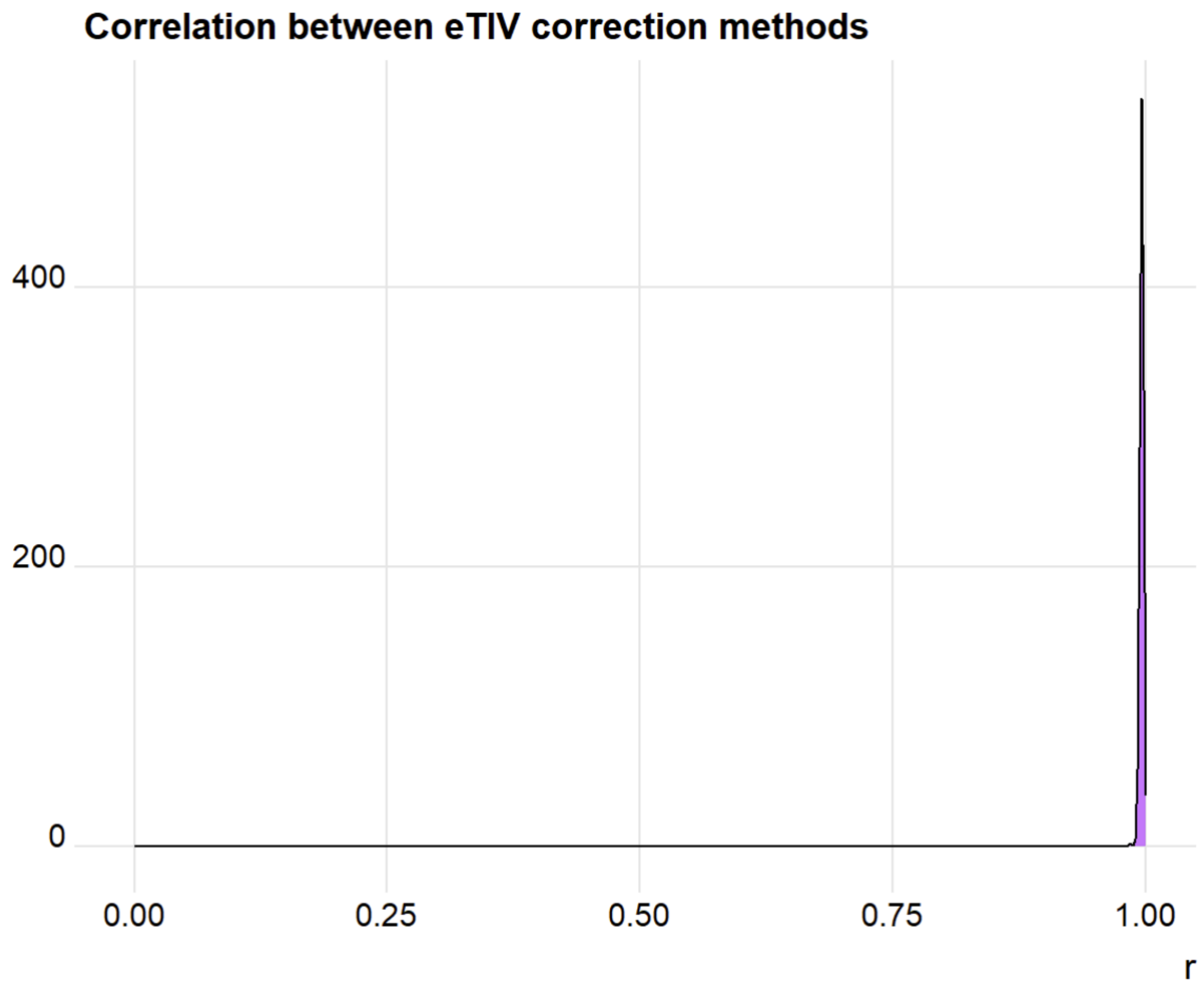


**Supplementary Figure S2. Different eTIV correction approaches converge.** Each brain feature was corrected for eTIV with two different methods, (1) using a residualisation approach, and (2) the power-corrected proportion method. For each feature, the results were correlated. The plot depicts the density across all correlation coefficients, indicating high convergence across the two tested approaches (minimum correlation r = 0.98).

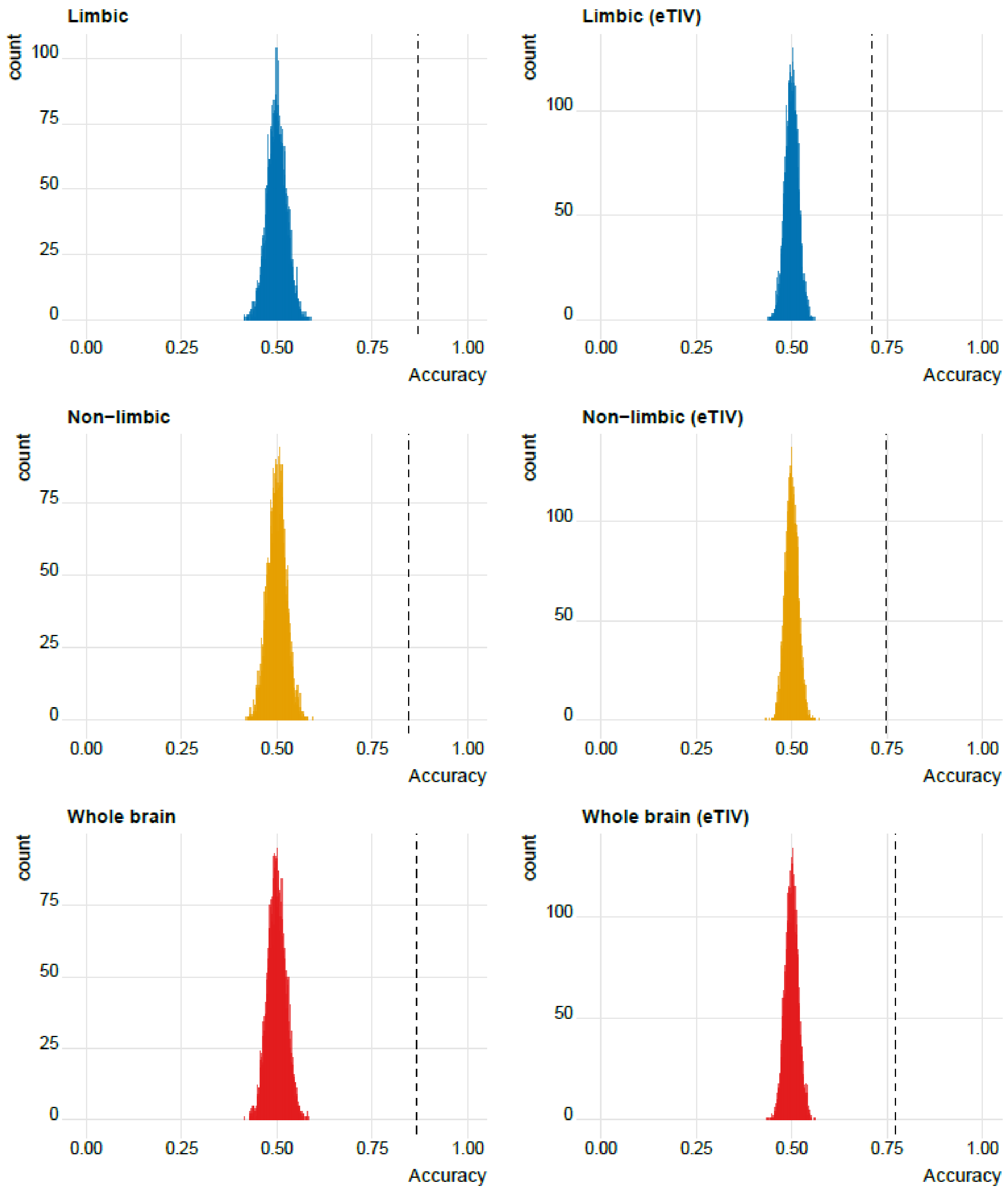


**Supplementary Figure S3. All model achieved significant accuracy.** No permutation-test based accuracy across 5000 permutations was higher than the accuracies achieved with the true model.

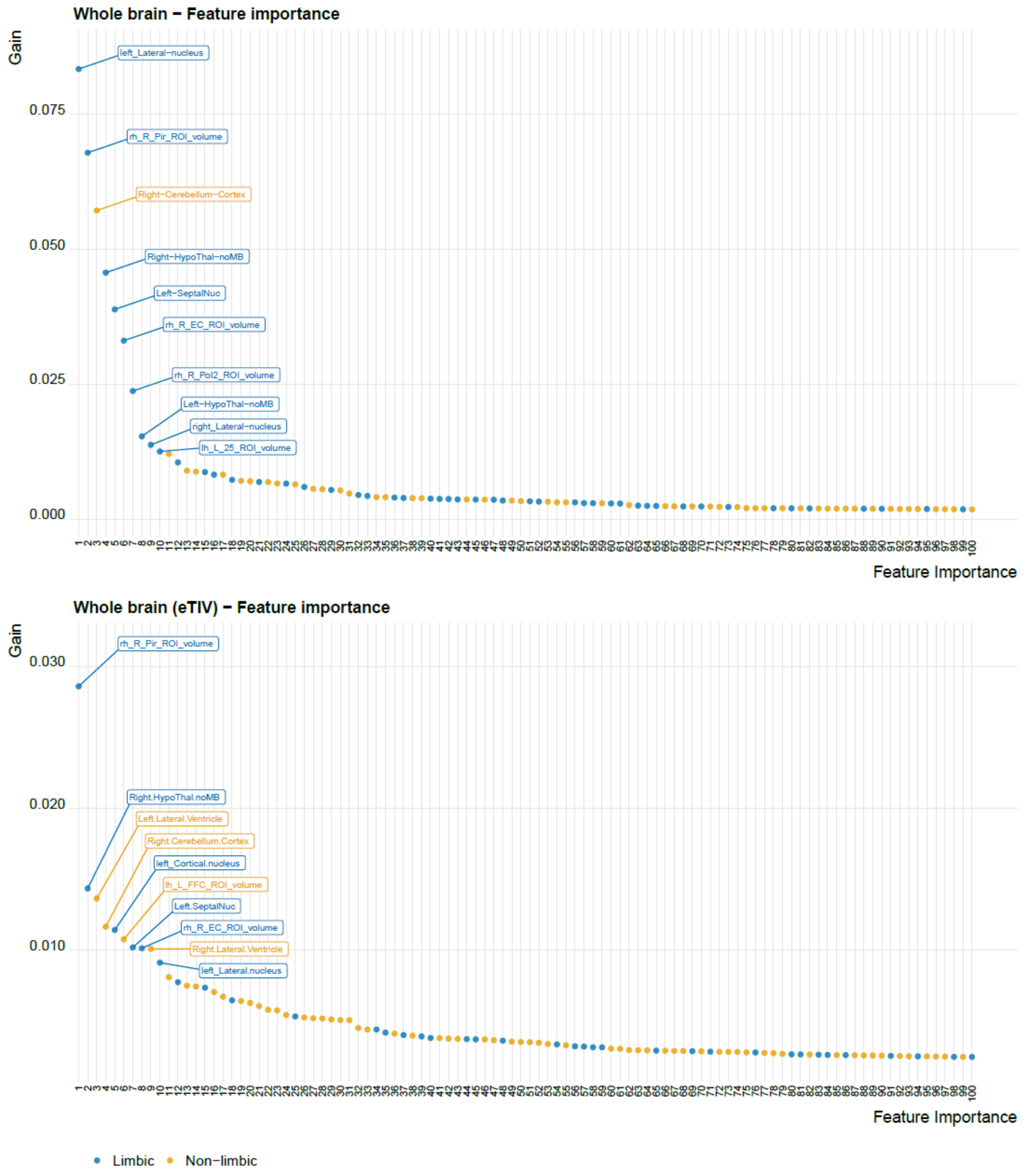


**Supplementary Figure S4. The majority of the 10 most important features of the whole brain models belong to the limbic system.** The feature importance of the first 100 features indicating the contribution of limbic and non-limbic structures.

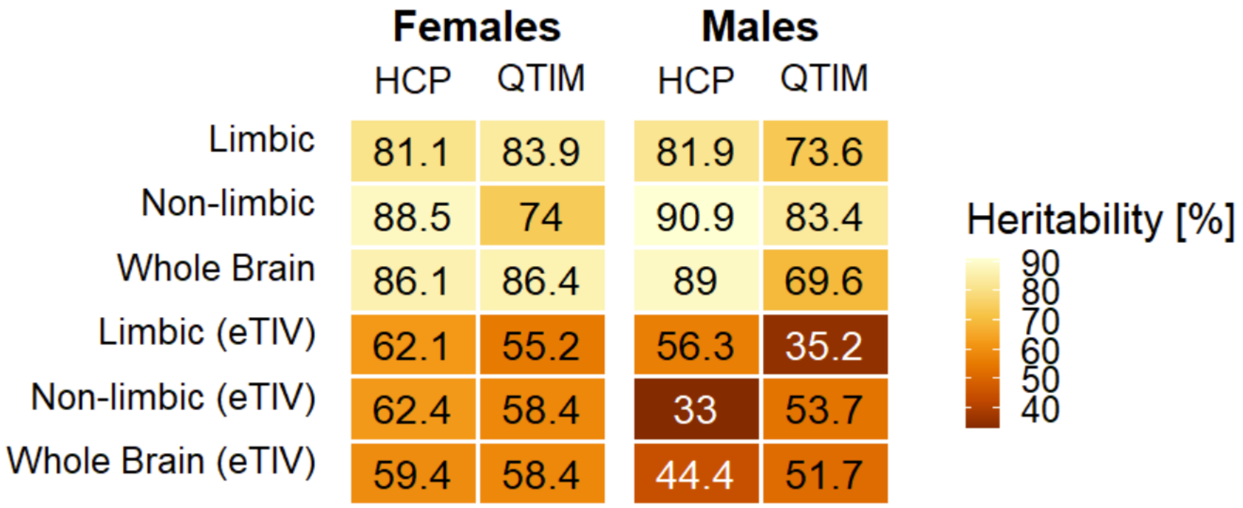


**Supplementary Figure S5. The class probabilities are heritable independently from sex.** Each cell shows the broad sense heritability in percent for females and males separately.

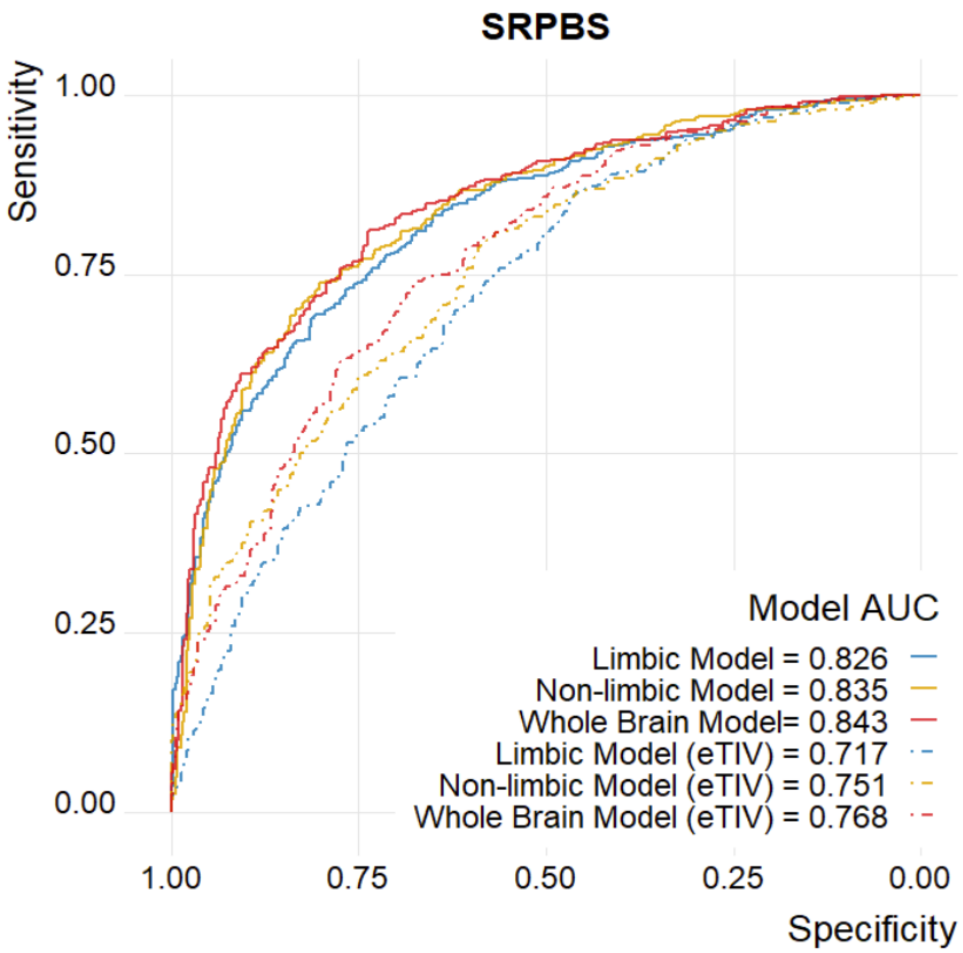


**Supplementary Figure S6. All models achieve high performance in the SRPBS sample.** The ROC curves show the performance of each model considering HC and MDD patients. The accuracies were 74.1%, 72.3%, and 73.1% for the limbic, non-limbic and whole brain model, respectively. When accounting for eTIV the accuracy decreased in all models, yielding 64.7%, 67.9% and 69.7% accuracy for limbic (eTIV), non-limbic (eTIV) and whole brain (eTIV), respectively.

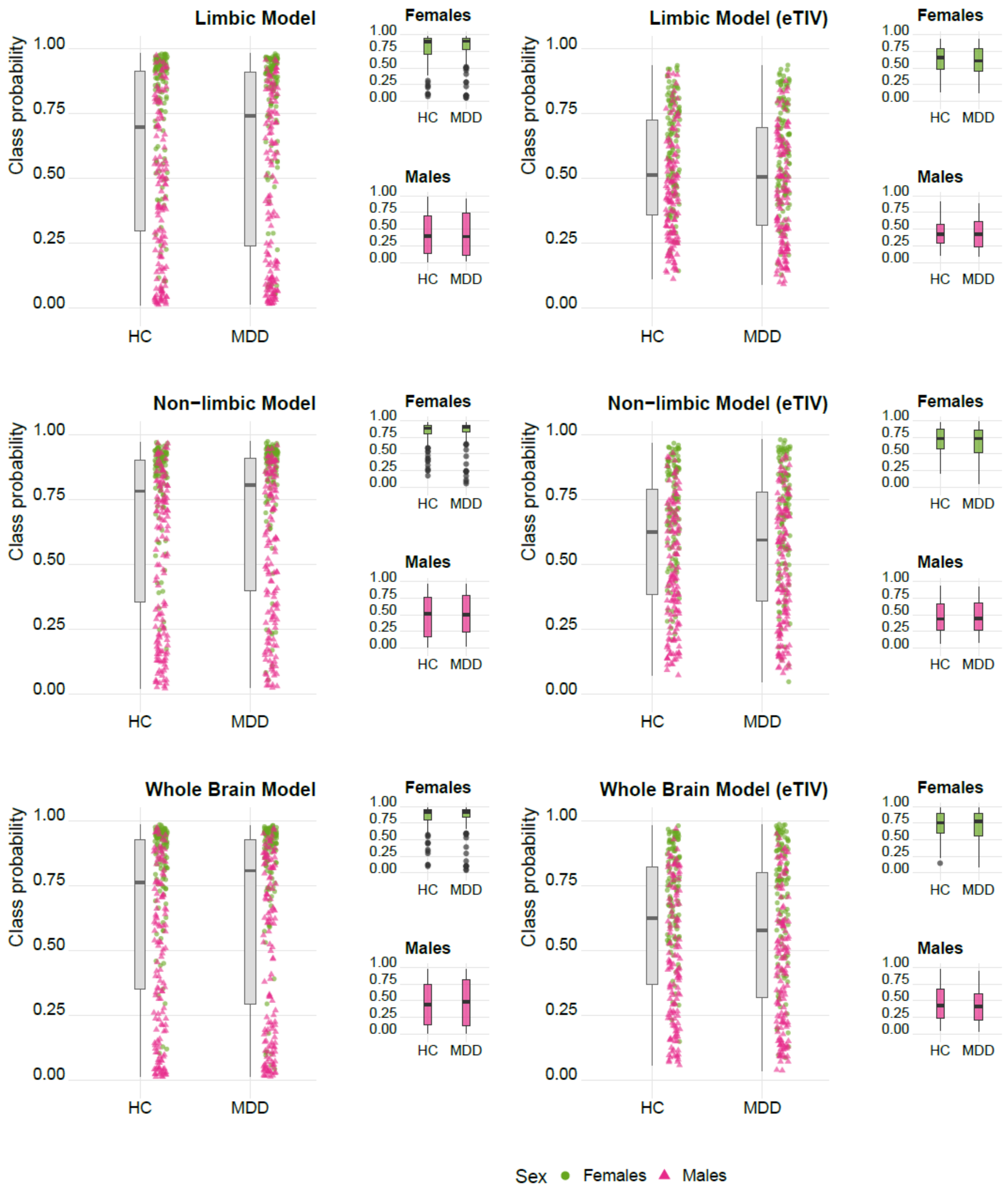


**Supplementary Figure S7. Limbic class probabilities in females are significantly higher in age-stratified analyses.** When matching healthy controls (HC) and Major Depressive Disorders patients (MDD) according to age, MDD diagnosis is significantly associated with higher limbic class probabilities in females. No significant effect was found for the other model or in males.

**Supplementary Table 1. List of limbic structures with the correspondent feature in Freesurfer**

| Structure | FreeSurfer Feature | Segmentation |
|---|---|---|
| | 33pr | |
| | p24pr | |
| | a24pr | |
| | p24 | |
| | a24 | |
| | p32pr | |
| | a32pr | |
| Anterior Cingulate Cortex | d32 | |
| | p32 | |
| | s32 | |
| | 8BM | |
| | 9m | |
| | 10v | Glasser atlas |
| | 10r | |
| | 25 | |
| Orbitofrontal Cortex | OFC | |
| | pOFC | |
| | MI | |
| | AVI | |
| | AAIC | |
| Insula | Ig | |
| | PI | |
| | PoI1 | |
| | PoI2 | |
| Piriform Cortex | Pir | |

| Structure | FreeSurfer Feature | Segmentation |
|---|---|---|
| Entorhinal Cortex | EC | |
| | PHA1 | |
| Parahippocampal Area | PHA2 | |
| | PHA3 | |
| | DVT | |
| | ProS | |
| | POS1 | |
| | POS2 | |
| | RSC | |
| | v23ab | |
| Posterior Cingulate Cortex | d23ab | |
| | 31pv | |
| | 31pd | |
| | 31a | |
| | 23d | |
| | 23c | |
| | PCV | |
| | Hippocampal_tail | |
| | subiculum-body | |
| | CA1-body | |
| | subiculum-head | |
| Hippocampus | hippocampal-fissure | Hippocampal Subfields |
| | presubiculum-head | |
| | CA1-head | |
| | presubiculum-body | |
| | parasubiculum | |

| Structure | FreeSurfer Feature | Segmentation |
| --- | --- | --- |
| | molecular_layer_HP-head | |
| | molecular_layer_HP-body | |
| | GC-ML-DG-head | |
| | CA3-body | |
| | GC-ML-DG-body | |
| | CA4-head | |
| | CA4-body | |
| | fimbria | |
| | CA3-head | |
| | HATA | |
| Amygdala | Lateral-nucleus | Nuclei of Amygdala |
| | Basal-nucleus | |
| | Accessory-Basal-nucleus | |
| | Anterior-amygdaloid-area-AAA | |
| | Central-nucleus | |
| | Medial-nucleus | |
| | Cortical-nucleus | |
| | Corticoamygdaloid-transitio | |
| | Paralaminar-nucleus | |
| Anterior and Dorsomedial Thalamic Nuclei | AV | Thalamic Nuclei |
| | LD | |
| | MDl | |
| | MDm | |
| Nucleus Accumbens | Nucleus-Accumbens | ScLimbic |
| Hypothalamus | HypoThal-noMB | |
| Fornix | Fornix | |

| **Structure** | **FreeSurfer Feature** | **Segmentation** |
|---|---|---|
| Mammillary Body | MammillaryBody | |
| Forebrain | Basal-Forebrain | |
| Septal Nuclei | SeptalNuc | |
